# Determinants of HIV infection among pregnant women in Cameroon: A contribution toward the elimination of vertical transmission in low- and middle-income countries

**DOI:** 10.1101/2024.05.15.24307434

**Authors:** Justin Ndié, Jean Pierre Yves Awono Noah, Francis Ateba Ndongo, Joseph Fokam, Alice Ketchaji, Rogacien Kana Dongmo, Christian Noël Bayiha, Richard Tchapda, Tatiana Avang Nkoa Palisson, Martial Gaël Bonyohe, Caroline Teh Monteh, Njamsnhi Yembe Wepnyu, Félicité Naah Tabala, Hernandez Lélé Siaka, Carelle Djofang Yepndo, Audrey Raïssa Djomo Nzaddi, Maurice Rocher Mbella, Marie Micheline Dongmo, Gildas Nguemkam, Ngo Issouck, Nelly Monkam, Leopoldine Madjo Oumbe, Clifford Moluh, Paul Tjek, Vittorio Colizzi, Carlo-Federico Perno, Giulia Cappelli, Nicaise Ndembi, David Kob, Gregory-Edie Halle Ekane, Basile Keugoung, Alexis Ndjolo, Serge Clotaire Billong, Céline Nkenfou, Jérôme Ateudjieu, Anne Cécile Zoung-Kanyi Bissek

## Abstract

**Background:** The risk of HIV transmission during antenatal care (ANC) in Cameroon remains a concern. According to recent studies, the prevalence of HIV in the country is around 4.5%, which increases the likelihood of vertical transmission.

**Objective:** To identify the determinants of HIV infection among pregnant women attending antenatal clinics (ANC) in Cameroon and to estimate HIV seroprevalence.

**Methods:** A cross-sectional study was conducted among ANC attendees aged ≥15 years from September 2022 to June 2023 in 324 health facilities in 08 regions of Cameroon (Adamaoua, Yaounde, East, Far-North, Douala, North, West, South). Sociodemographic and clinical data were collected using questionnaire. HIV screening was performed according to the national algorithm. Estimates of HIV seroprevalence and identification of its determinants using multivariable logistic regression (95% CI) were performed with Excel and SPSS 22 software.

**Results:** Overall, 10674 pregnant women were enrolled, with median [IQR] age 25 years [21– 30]; 40.0% at a secondary educational level; 44.1% married monogamously; 46.3% multiparous; 38.8% in the second quarter of pregnancy and 16.5% reporting at least one abortion. Overall HIV seroprevalence was 2.6% [95%CI: 2.33; 2.93]. Significantly higher prevalence was found with the regions of Adamaoua (aOR 3.78 [95%CI: 1.87-7.67], p<0.001), East (9.38 [5.6-15.67], p<0.001), North (3.07 [1.74-5.42], p<0.001), South (2.93 [1.66-5.16]; p<0.001); lack of education (2.08 [1.06-4.06], p=0.032), primary education (2.44 [1.32-4.50], p=0.004) and secondary education (2.29 [1.28-4.08], p=0.005) were significantly associated with HIV infection, while monogamous marriage (0.33 [0.22-0.51], p<0.001), the absence of abortion (0.59 [0.37-0.98], p=0.036) and large multiparous (0.38 [0.17-0.82]; p=0.015) were protective.

**Conclusion:** Despite the overall low-prevalence among pregnant women at national-level, several factors are associated with HIV in ANC, the absence or low-level of education, being elderly (>30 years), singleness, history of abortion and low parity predicted the HIV status during ANC. Thus, public health interventions towards these at-risk target groups will help to reduce new infections among pregnant women, hence contributing to achieve eMTCT in Cameroon.

## Introduction

HIV infection is still prevalent in pregnant women and remains a major public health concern across several low-and middle-income countries (LMICs). Of note, about 1.5 million pregnant women are living with HIV worldwide (1,2), sub-Saharan Africa is home to two-third of cases worldwide (3), with prevalence varying between 5% and 42% among pregnant women (4).

Pregnant women living with HIV are at high risk of vertical transmission to their infants during pregnancy, childbirth or breastfeeding (5). Of relevance, in the absence of any intervention, between 20% and 45% of infants could contract HIV, with an estimated risk of 5-10% during pregnancy, 0-20% during labour and delivery and 5-20% during breastfeeding (4–9). Furthermore, without treatment, half of all HIV-infected children will die before their second birthday, thereby stressing the need to consider the elimination of HIV vertical in high priority countries, including Cameroon (4).

Despite a 50% decline in HIV prevalence from 5.5% in 2005 (10) to 2.7% in 2018 (11) in the general population, the burdens of HIV in specific populations including pregnant women remain high in Cameroon. Although HIV prevalence among pregnant women fell from 7.6% in 2009 to 4.26% in 2019 in Cameroon, some geographical regions still stand at higher risks (i.e. South with 8.46%), underscoring the need for regular surveillance to ensure evidence-based public health actions for eliminating HIV vertical transmission (12). Furthermore, HIV prevalence among pregnant women in Cameroon is similarly distributed, ranging from 5.58% in urban areas to 5.87% in rural settings according to reports of the sentinel surveillance survey of 2019 (13,14).

Several factors contribute to the dynamics of this infection in pregnant women in these Cameroonian settings, among which being single as marital status, multiparity, age and living in specific regions were significantly associated risk of contracting HIV infection (14). Thus, controlling these drivers of HIV infection among pregnant women is an essential component in achieving the global effort in eliminating paediatric HIV by 2030. However, with population dynamics and behavioural changes overtime, it is of paramount importance to generate update evidence for strategic information toward priority interventions at the public health level. Therefore, our study objective was to update HIV epidemiology and identify determinants of infection among pregnant women attending antenatal clinics in Cameroon.

## Methods and materials

### Ethical considerations

Ethical approval was obtained from the National Ethics Committee for Human Health Research (reference number **N°2022/08/1478/CE/CNERSH/SP**) and an Administrative Research Authorization (reference number **N° 631-26-22**) was obtained. Written informed consent was obtained from study participants. Furthermore, parental or guardian consent has been obtained for each pregnant woman under the age of 21. It should be noted that the legal age of majority in Cameroon is 21.

### Study design

A national cross-sectional study was carried out between September 2022 and June 2023 among pregnant women receiving antenatal care (ANC) in 324 facilities in eight regions of Cameroon: Adamaoua, East, Far North, North, West, South, Yaounde and Douala. Yaounde and Douala are major cities comparable to other regions of the country due to their demographic weight. The health facilities were selected on the basis of their monthly ANC weight.

### Study population

The study population consisted of pregnant women aged 15 and over, who consented to participate in the study and were receiving antenatal and maternity care. In addition, they had to be of unknown HIV status or have been declared HIV-negative for at least three months. Pregnant women receiving antiretroviral treatment and/or presenting with an acute illness likely to interfere with the study process were not included.

### Sample size and sampling

The sample size was calculated using the WHO expert formula (Adequacy of sample size in health surveys) and taking into account the HIV prevalence of pregnant women observed in previous studies at national level (13,15). Accordingly, the sample size was estimated at 10573 pregnant women.

### Data collection and HIV testing procedure

Two HIV tests were used according to national guidelines (national HIV screening algorithm): Determine (Abbott Laboratories, IL, USA) and Oraquick.

Data collection was carried out by health workers (Nurses and Midwives) in the antenatal and maternity consultation departments of the health facilities selected for the study. They were trained for 03 days in the use of Determine and Oraquick for HIV screening and in the study methodology. Testing took place at the entry point (antenatal and maternity clinics) of the health facilities selected for the study. The number of pregnant women to be recruited at each health facility was based on the average number of antepartum consultations per month. Pregnant women were recruited by voluntary sample until the size of the workforce per site was reached. When a pregnant women arrived at the ANC or maternity ward, the site’s trained provider introduced himself to her, checked her eligibility, sent her the information leaflet and asked for her informed consent.

Once the participants had provided informed consent, pre-test counselling was conducted, after which HIV testing of pregnant women was performed in accordance with the national algorithm (Figure 1). Pregnant women who tested positive by the national HIV screening algorithm were managed by the aforementioned health facility in accordance with the guidelines for HIV management in Cameroon.

**Figure 1:**
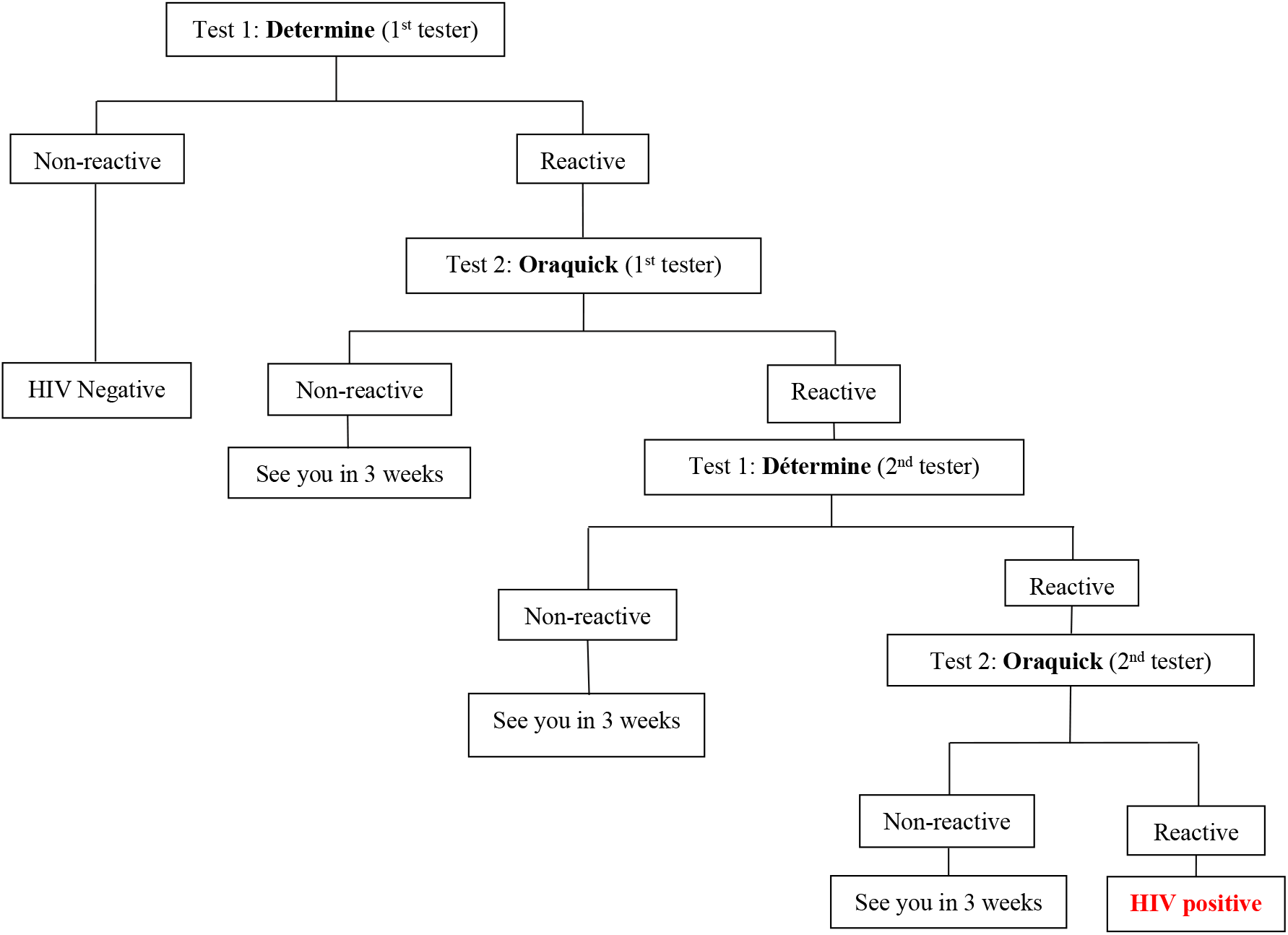
HIV screening algorithm used in the study

### Data management

Data were collected on a specific register developed for the purpose of the study. Collected data were then entered using CS Pro 7.7.3 and analysed using SPSS 22. Numerical variables were described using the median with the interquartile range (IQR), and categorical variables were described using proportions. Multivariate logistic regression was used to identify the determinants of HIV seroprevalence. The variables included in the multivariate regression were: region, age, level of education, marital status, age of pregnancy, number of pregnancies and history of abortions. Indeterminate results interpreted by healthcare staff were excluded from the model. ***HIV seroprevalence*** was defined as the proportion of pregnant women who tested positive by the HIV screening algorithm.

## Results

### Sociodemographic characteristics of pregnant women

Of 10,673 pregnant women tested, the median age was 25 (21 - 30) and the age group most represented was 25-30 (31.3%). 44.1% were in a monogamous marriage, 46.3% of pregnant women were multiparous, 38.8% of pregnant women were in the 2nd trimester of pregnancy and 16.5% of pregnant women had declared that they had already had at least one abortion.

**Table I:** Sociodemographic characteristics of pregnant women.

| Sociodemographic characteristics | N= 10,673 |  |
| --- | --- | --- |
| Region | not | % |
| *Adamaoua* | 740 | 6.9 |
| *East* | 838 | 7.9 |
| *Far North* | 2091 | 19.6 |
| *North* | 2054 | 19.2 |
| *West* | 110 | 1.0 |
| *South* | 732 | 6.9 |
| *Yaounde* | 2173 | 20.4 |
| *Douala* | 1935 | 18.1 |
| **Median age (IQR)** | 25 years (21 – 30) |  |
| *Under 20* | 1687 | 15.8 |
| *20 - 25 years old* | 3121 | 29.3 |
| *25 - 30 years old* | 3336 | 31.3 |
| *30 - 35 years old* | 1649 | 15.5 |
| *Over 35 years old* | 859 | 8.1 |
| **Educational level** |  |  |
| *Never schooled* | 2149 | 20.6 |
| *Primary* | 2729 | 26.2 |
| *Secondary 1st cycle* | 2644 | 25.3 |
| *Secondary 2nd cycle* | 1529 | 14.7 |
| *Superior* | 1384 | 13.3 |
| **Marital status** |  |  |
| *Monogamous bride* | 4679 | 44.1 |
| *Polygamous Bride* | 1474 | 13.9 |
| *Cohabitation* | 2325 | 21.9 |
| *Bachelor* | 2132 | 20.1 |
| **Number of pregnancies** |  |  |
| *Nulliparous* | 3160 | 30.9 |
| *Primiparous* | 2344 | 22.9 |
| *Pauciparous* | 3019 | 29.5 |
| *Multiparous* | 1184 | 11.6 |
| *Large multiparous* | 528 | 5.2 |
| **Gestational age (in weeks of amenorrhea)** | 24 weeks (16 – 32) |  |
| *1st trimester of pregnancy* | 1452 | 14.1 |
| *2nd trimester of pregnancy* | 3986 | 38.8 |
| *3rd trimester of pregnancy* | 4835 | 47.1 |
| **History of abortion** |  |  |
| *No abortion* | 8478 | 83.5 |
| *01 abortion* | 1105 | 10.9 |
| *02 abortions and more* | 566 | 5.6 |

#### Seroprevalence of HIV infection among pregnant women

Overall, HIV seroprevalence among pregnant women was 2.6%, 95% CI [2.33; 2.93].

**Table II:**
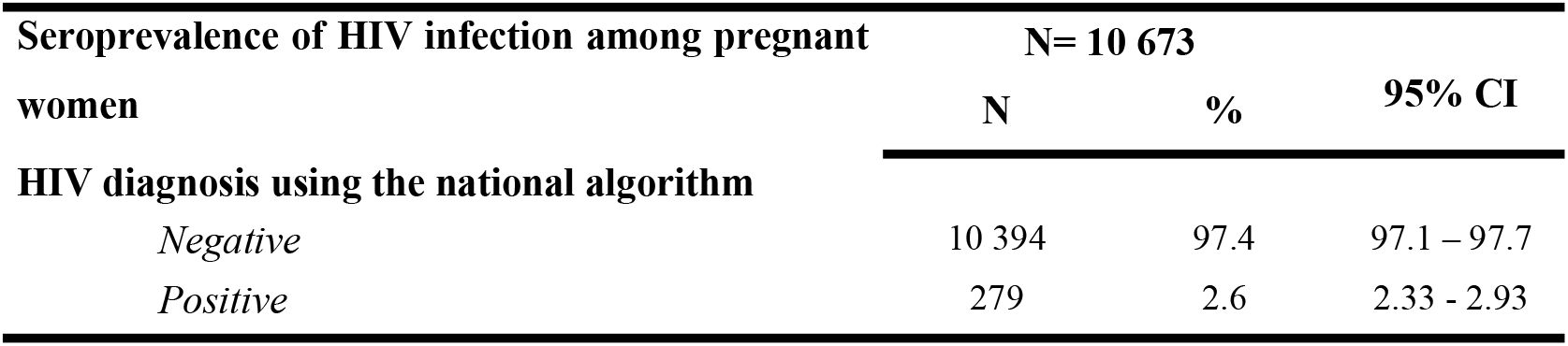
Seroprevalence of HIV infection among pregnant women.

### Factors associated with HIV seroprevalence among pregnant women

The regions of Adamaoua (aOR (95% CI): (3.78 (1.87-7.67); p<0.001), East (9.38 (5.6-15.67); p< 0.001), North 3.07 (1.74-5.42); p<0.001), South (2.93 (1.66-5.16); p<0.001), non-schooling in the pregnant women (2 .08 (1.06-4.06); p=0.032), the level of primary education (2.44 (1.32-4.50); p=0.004), the level of secondary education (2 .29 (1.28-4.08); p=0.005), nulliparity (2.67 (1.21-5.87); p=0.015), primiparity (2.72 (1.27-5 .79); p=0.010), pauciparity (2.63 (1.27-5.79); p=0.007) and multiparity (2.14 (1.03-4.45); p=0.041) were significantly associated with HIV infection. On the other hand, monogamous marriage (0.33 (0.22-0.51); p<0.001), the absence of notion of abortion among the pregnant women (0.59 (0.37-0.98); p=0.036) were associated with a reduced risk of HIV infection.

**Table III:** Determinants of seroprevalence of HIV infection among pregnant women

|  | HIV diagnosis |  | Univariable analysis |  |  | Multivariate analysis |  |  |
| --- | --- | --- | --- | --- | --- | --- | --- | --- |
|  | Negative | Positive | B(SE) | P | OR(95% CI) | B( SE ) | P | aOR (95% CI) |
| **Region** |  |  |  |  |  |  |  |  |
| Adamaoua | 79.7% | 2.3% | 0.471(0.19) | 0.129 | 1.6(0.87;2.94) | **1.33\*\*\* (0.36)** | **0.001** | **3.78(1.87;7.67)** |
| East | 91.2% | 8.8% | **1.89\*\*\* (0.23)** | **0.001** | **6.59(4.24; 10.27)** | **2.238\*\*\* (0.26)** | **0.001** | **9.38(5.6;15.66)** |
| Far North | 97.8% | 2.2% | 0.45(0.24) | 0.063 | 1.57(0.98;2.51) | **1.16\*\*\* (0.31)** | **0.001** | **3.2(1.74;5.84)** |
| North | 97.6% | 2.4% | **0.51\* (0.24)** | **0.033** | **1.66(1.04;2.66)** | **1.121\*\*\* (0.29)** | **0.001** | **3.07(1.74;5.42)** |
| West | 97.3% | 2.7% | 0.64(0.62) | 0.293 | 1.91(0.57, 6.38) | 0.834(0.76) | 0.270 | 2.30(0.52;10.12) |
| South | 96.0% | 4.0% | **1.03\*\*\* (0.27)** | **0.001** | **2.81(1.66;4.76)** | **1.073\*\*\* (0.29)** | **0.0001** | **2.93(1.66;5.16)** |
| Yaounde | 98.5% | 1.5% | 0.18(0.26) | 0.946 | 1.018(0.61;1.69) | 0.029(0.27) | 0.917 | 1.03(0.60, 1.76) |
| Douala | 98.6% | 1.4% |  |  | 1 |  |  | 1 |
| **Age** |  |  |  |  |  |  |  |  |
| Under 20 | 97.3% | 2.7% | -0.278(0.186) | 0.246 | 0.76(0.47;1.21) | **-1.192\*\*\* (0.32)** | **0.001** | **0.30(0.16;0.57)** |
| 20 - 25 years old | 97.8% | 2.2% | **-0.485\* (0.223)** | **0.029** | **0.615(0.39;0.95)** | **-1.083\*\*\* (0.28)** | **0.001** | **0.34(0.19, 0.58)** |
| 25 - 30 years old | 97.8% | 2.2% | **-0.481\* (0.220)** | **0.029** | **0.618(0.40, 0.95)** | **-0.822\*\*\* (0.25)** | **0.001** | **0.44(0.27;0.71)** |
| 30 - 35 years old | 96.3% | 3.7% | 0.06(0.227) | 0.793 | 1.06(0.68, 1.66) | -0.058(0.25) | 0.814 | 0.94(0.58, 1.82) |
| Over 35 years old | 96.5% | 3.5% |  |  | 1 |  |  | 1 |
| **Level of education** |  |  |  |  |  |  |  |  |
| Out of school | 97.5% | 2.5% | **0.79\*\* (0.29)** | **0.006** | **2.20(1.25; 3.87)** | **0.732\* (0.34)** | **0.032** | **2.08(1.06;4.06)** |
| Primary | 96.8% | 3.2% | **1.02\*\*\* (0.27)** | **0.001** | **2.78(1.63;4.76)** | **0.892\* (0.31)** | **0.004** | **2.44(1.32;4.50)** |
| Lower secondary | 96.6% | 3.4% | **1.10(0.27)** | **0.001** | **3.01(1.76;5.15)** | **0.828\* (0.29)** | **0.005** | **2.29(1.28;4.08)** |
| Secondary <sup>2nd</sup> cycle | 98.4% | 1.6% | 0.31(0.25) | 0.34 | 1.36(0.72;2.57) | 0.130(0.34) | 0.704 | 1.14(0.58;2.22) |
| Superior | 98.8% | 1.2% |  |  | 1 |  |  | 1 |
| **Marital status** |  |  |  |  |  |  |  |  |
| Monogamous bride | 98.0% | 2.0% | **-0.489\*\* (0.122)** | **0.002** | **0.613(0.45, 0.84)** | **-1.094\*\*\* (0.21)** | **0.001** | **0.33(0.22, 0.51)** |
| Polygamous Bride | 97.4% | 2.6% | -0.208(0.203) | 0.307 | 0.813(0.55, 1.21) | **-0.969\*\*\* (0.26)** | **0.001** | **0.38(0.23, 0.64)** |
| Cohabitation | 96.7% | 3.3% | -0.10(0.169) | 0.951 | 1.01(0.72, 1.41) | **-0.441\* (0.19)** | **0.021** | **0.64(0.44, 0.94)** |
| Bachelor | 96.8% | 3.2% |  |  | 1 |  |  | 1 |
| **Gestational age** |  |  |  |  |  |  |  |  |
| 1st trimester of pregnancy | 96.8% | 3.2% | 0.331(0.178) | 0.063 | 1.39(0.98, 1.97) | **0.559\* (0.19)** | **0.004** | **1.75(1.19;2.57)** |
| 2nd trimester of pregnancy | 97.1% | 2.9% | 0.226(0.135) | 0.095 | 1.25(0.96; 1.63) | **0.311\* (0.15)** | **0.034** | **1.37(1.02;1.82)** |
| 3rd trimester of pregnancy | 97.7% | 2.3% |  |  | 1 |  |  | 1 |
| **Parity** |  |  |  |  |  |  |  |  |
| Primiparous | 97.7% | 2.3% | 0.033(0.322) | 0.919 | 1.03(0.55, 1.94) | 0.017(0.20) | 0.93 | 1.02(0.68;1.52) |
| Pauciparous | 97.1% | 2.9% | 0.244(0.312) | 0.434 | 1.28(0.69;2.35) | -0.014(0.21) | 0.95 | 0.99(0.65;1.49) |
| Multiparous | 96.9% | 3.1% | 0.327(0.327) | 0.331 | 1.39(0.12;2.68) | -0.22(0.27) | 0.42 | 0.80(0.47; 1.36) |
| *Large multiparous* | 97.7% | 2.3% | 0.33(0.34) | 0.33 | 1.39(0.72;2.68) | **-0.98\*(0.40)** | **0.015** | **0.38(0.17;0.82)** |
| *Nulliparous* | 97.8% | 2.2% |  |  | 1 |  |  | 1 |
| **History of abortion** |  |  |  |  |  |  |  |  |
| *No abortion* | 97.6% | 2.4% | **-0.485\*(0.229)** | **0.034** | **0.62(0.39, 0.96)** | **-0.519\*(25)** | **0.036** | **0.59(0.37, 0.98)** |
| *01 abortion* | 97.2% | 2.8% | -0.337(0.28) | 0.225 | 0.71(0.41, 1.25) | -0.259(0.29) | 0.379 | 0.77(0.43, 1.37) |
| *02 abortions and more* | 96.1% | 3.9% |  |  | 1 |  |  | 1 |

1: reference category; **B(ES)**: regression coefficient (standard error); **P**: significance; \*: significant at 5%; \*\*: significant at 1%; **aOR**: adjusted
Odds Ratio

Regions aOR [CI 95%] Adamaoua 3.78 [1.87-7.67]; p<0.001, East (9.38 [5.6-15.67] ; p<0.001, North 3.07 [1.74-5.42]; p<0.001, South 2.93 [1.66-5.16]; p<0.001, non-enrolment among the pregnant women 2.08 [1.06-4.06]; p=0.032, primary education level 2.44 [1.32-4.50]; p=0.004 and secondary education 2.29 [1.28-4.08]; p=0.005 were significantly associated with HIV infection. On the other hand, monogamous marriage status 0.33 [0.22-0.51]; p<0.001, the absence of any notion of abortion in the pregnant women 0.59 [0.37-0.98]; p=0.036 and large multiparous 0.38 [0.17-0.82]; p=0.015 were associated with a reduced risk of HIV infection.

## Discussion

Evidence on HIV infection and its predictors among pregnant women is key to ensuring an HIV free new generation of children beyond 2030. Most importantly, LMICs like Cameroon require such epidemiological surveillance to set-up priority interventions with impact at country-level. In the present study, HIV seroprevalence among pregnant women was 2.6% IC95% [2.33; 2.93]. These results are similar to the last national HIV prevalence rate (2.7%) conducted in 2018 (11). Similarly, our findings are in close alignment with those of previous studies conducted in Burkina Faso (2.16%) and Mali (2.85%) (16,17). The present prevalence is significantly lower compared to the 2016 and 2019 sentinel surveys with 5.75% and 4.26% respectively among pregnant women (12,13), including the results of studies which reported HIV prevalences of 5.70% (95% CI: 4.93-6.40), 6% (3.0-10.2%) and 7.8% respectively among pregnant women in Cameroon (18–20). Furthermore, the respective prevalence rates of 9.2% and 7.22% reported in the countries of Chad and Nigeria are also of relevance (21,22). Our findings therefore reflect the declining trend of HIV among pregnant women in Cameroon (7.6% in 2009 to 2.9% in 2016 (19,23) and from 3.4% in 2018 to 2.1% in 2022 (24–26). This declining prevalence is the result of progress and strategies implemented by Cameroon in recent years to strengthen HIV prevention among women, who are disproportionately affected by HIV. These include the integration of reproductive health and maternal, newborn, child and adolescent health services/HIV/PMTCT, decentralisation of services and delegation of tasks, family-based HIV testing, implementation of option B+, contact tracing, implementation of Users Fees and HIV self-testing for partners of pregnant women (24).

The spatial distribution of HIV among pregnant women shows regional disparities varying from 2.2% in the Far North to 8.8% in the East and similarly low prevalence in the country major cities of Yaoundé (1.5%) and Douala (1.4%) (11). In similar surveys, regional variations from 0.7% in the Far North to 11.8% in the South, as well varying trends between urban and rural settings (14,19). These regional and urban disparities show that prevention activities and priority interventions achieved the expected goals more easily in the major cities, likely due to accessibility to several channels and means of information and communication. In addition, the educational level and the presence of community-based organisations in these major cities also contribute to strengthening HIV prevention, raising awareness and involving pregnant women and their partners in healthy behavioural factors. Henceforth, these results suggest strengthening strategies through targeted and differentiated priority HIV prevention interventions in regions, also supported by previous the Demographic and Health Survey (11).

Regarding drivers of HIV infection, single pregnant women were more likely to be infected with HIV than married or cohabiting women. These results are similar to previous reports (14,27), likely due to the fact that unmarried women are more likely to have several sexual partners thus increasing the risk of infection. Also, their economic vulnerability of most single women also exposes them to transactional sex (28). In this study, the risk of HIV infection increased with age. Indeed, pregnant women aged 30 and over were more likely to be infected with HIV than younger women. These results are comparable to those reported in the study by Anoubissi and al (14). These results are consistent with the distribution of HIV infection in the general population and suggest that younger women are better at implementing HIV prevention measures than older women. Lower level of education was significantly associated with a two-folds risk of acquiring HIV infection, as previously reported similarly reported in Cameroon and elsewhere (18,29). Thus, HIV prevention should prioritise women with low educational level, as they appear to poorly assimilate and implement HIV preventive measures. Gestational age, parity and history of abortion were significantly associated with HIV infection. Indeed, pregnant women in their 1^st^ or 2^nd^ trimester were almost 2 times more likely to be infected with HIV than those in their 3^rd^ trimester. Similarly, nulliparous, primiparous, pauciparous or multiparous pregnant women were almost 2 times more likely to be infected with HIV than large multiparous women. While suggesting further investigation on this area, Rindcy Davis *et al* showed that primiparous women were associated with a slightly lower prevalence of HIV testing (aPR 0.97, 95% CI 0.95, 0.99) than nulliparous women (30). Furthermore, pregnant women who had no abortions were 41% less likely to be infected with HIV than those who had had two or more abortions. Abortions generally traumatise and cause cervical and vaginal lesions, which probably increase the risk of contracting STIs, including HIV.

The potential limitations of this study would be the inability to detect cases of early infection, which would have been handled by performing nucleic acid testing or p24 antigen detection. However, this might have limited inference on the present evidence.

## Conclusion

Despite the overall low-prevalence among pregnant women at national-level (2.6%), there are target population still experiencing higher risk of infection within the frame of an ongoing pregnancy, which include the absence or low-level of education, being elderly (>30 years), singleness, history of abortion and reporting low parity during ANC. Thus, public-health interventions towards these at-risk target groups will further limit events of new HIV infections among pregnant women, hence contributing to achieve eMTCT in Cameroon and other LMICs sharing similar epidemiological and programmatic features.

## Data Availability

The data are available

## Acknowledgments

We would like to thank all the participants in the study for their contributions. We would also like to express our gratitude to Dr. Bacha Abdelkader for his collaboration and commitment in the implementation of the study. We would also like to thank the Cameroon Ministry of Public Health, UNICEF Cameroon and the Health Project Implementation Unit of the Islamic Development Bank for providing the necessary and accurate resources.

